# Outpatient screening of health status and lifestyle among post-bariatric patients during the Covid-19 pandemic in Sao Paulo, Brazil.

**DOI:** 10.1101/2020.07.30.20165068

**Authors:** Karla Fabiana Goessler, Carolina Ferreira Nicoletti, Diego Augusto Nunes Rezende, Sofia Mendes Sieczkowska, Gabriel Perri Esteves, Rafael Genario, Gersiel Nascimento de Oliveira Júnior, Kamila Meireles, Ana Jéssica Pinto, Michele Nakahara-Melo, Roberto de Cleva, Marco Aurélio Santo, John Kirwan, Hamilton Roschel, Bruno Gualano

**Affiliations:** Applied Physiology and Nutrition Research Group, School of Physical Education and Sport; Faculdade de Medicina FMUSP, Universidade de São Paulo, São Paulo, SP, BR, University of Sao Paulo, SP, BR; Departamento de Ciências da Saúde/Departamento de Clínica Médica, Faculdade de Medicina de Ribeirão Preto, Universidade de São Paulo, Ribeirão Preto, SP, BR, University of Sao Paulo, SP, BR; Programa de Pós-Graduação em Enfermagem na Saúde do Adulto, Escola de Enfermagem, Universidade de São Paulo, São Paulo, SP, BR, University of Sao Paulo, SP, BR; Department of Digestive Surgery, School of Medicine, University of Sao Paulo; Integrated Physiology and Molecular Medicine Laboratory, Pennington Biomedical Research Center, Baton Rouge, LA, USA; Rheumatology Division, Hospital das Clinicas HCFMUSP, Faculdade de Medicina,Universidade de São Paulo

**Keywords:** bariatric surgery, obesity, health care, social isolation, SARS-CoV-2

## Abstract

**Background/Objectives:** This was an out-of-hospital screening of health status and lifestyle during the Covid-19 pandemic in post-operative bariatric patients from Sao Paulo, Brazil, prevented from face-to-face health care.

**Subjects/Methods:** In this cross-sectional study, 66 patients were remotely (via phone call) and in-person (by home visit) assessed for health status and lifestyle habits. Results: Mean age was 47.4 years. Patients were obese grade I (30.0%), II (22.0%), and III (30.0%), and 94.2% had above reference waist circumference values. Sixty-four percent displayed high blood pressure, whereas 24% showed CRP levels above normal range. Nineteen percent of patients reported irregular use of nutritional supplementation and 6.0% reported binge eating habits. Thirty-three exhibited symptoms of depression. Mild-to-moderate and moderate-to-severe anxiety symptoms were reported by 27.4% and 11.3% of the patients; 4.5% exhibited suicidal ideation and were referred to a specialist for healthcare. Of relevance, inactive patients (59.6%) had poorer global mental and physical health scores as compared to active peers (both p<0.05). Conclusion: This out-of-hospital screening revealed that the absence of face-to-face health care due to the Covid-19 pandemic is associated with suboptimal status of physical and mental health as well as lifestyle inadequacies among patients who have recently undergone bariatric surgery.

**Key points:**

- We performed an out-of-hospital screening in post-operative bariatric patients prevented from face-to-face health care during the Covid-19 pandemic.
- Sixty-five percent displayed high blood pressure, whereas 24% showed C-reactive protein levels above normal range.
- About one third showed mild to severe symptoms of depression, whereas ∼40% showed mild to severe anxiety symptoms.
- Inactive patients (59.6%) had poorer global mental and physical health scores as compared to active peers.
- Three patients exhibited suicidal ideation and were referred to a specialist for healthcare.
- During the Covid-19 pandemic, there are a considerable number of post-bariatric patients in need of direct health care.

---

As of July 30, Brazil counted over 2,556,207 cases and over 90,000 deaths due to Covid-19, with disease spread continuing unabated. On March 24, the government of Sao Paulo, the epicenter of the pandemic in Latin America, issued a set of measures for social distancing (including a stay-at-home order) to contain new cases and mitigate virus contagion. Simultaneously, COVID-19 is placing enormous pressure on public health systems across the globe. The Clinical Hospital in the School of Medicine at the University of Sao Paulo, the largest quaternary hospital in Latin America, responded to the outbreak by reassigning 1,000 beds exclusively for patients diagnosed with Covid-19. As a result, however, elective medical appointments had to be temporarily suspended, potentially predisposing patients with complex conditions to deterioration in overall health and quality of life.

Out of concern that COVID-19 has created a situation where at-risk patients are confined with little or no access to face-to-face health care, we performed an out-of-hospital screening of health status and lifestyle habits remotely (by phone calls and/or visiting at home) among patients who recently underwent bariatric surgery, a condition that requires systematic health care follow-up to ensure patient safety and to sustain the benefits of the surgical procedure ^1,2,3^. Data were collected between April 20 and July 9, 2020, a period in which social distancing measures were in place in the city of Sao Paulo, Brazil. This study was approved by the local ethical committee, and patients provided written informed consent before participation.

We assessed post-bariatric patients from the outpatient Bariatric Surgery Clinic, in the Clinical Hospital at the University of Sao Paulo. Eligible patients were ≥ 18 years old, with a surgery elapsed time ≤ 12 months, and without any COVID-19 symptoms.

The 66 participants received one home visit and/or three phone calls for interview. Twelve patients (18.2%) were interviewed by phone calls only as they lived outside Sao Paulo city. During the visit, patients completed a demographic questionnaire that included information about age, sex, self-reported use of medication and comorbidities. Following a 10-min rest, blood pressure was measured three times using an automated device (Omron, HEM-7122). Fasting blood samples were collected to analyze glucose, lipid profile, and C-reactive protein (CRP). Anthropometric measures (waist circumference, weight and height) were also performed ^4^. Reference values for cardiometabolic risk factors were: ≤120 mmHg and/or ≤80 mmHg for systolic and diastolic blood pressure ^5^; <80 cm and <94 cm for waist circumference for men and women ^6^, respectively, ≤99 mg/dL for fasting glucose level; <150 mg/dL, >40 mg/dL, ≤129 mg/dL, and <190 mg/dL for triglycerides, HDL, LDL, and total cholesterol, respectively; <5.0 mg/dL for CRP. The references values for blood biochemistry were provided by the Central Laboratory of the Clinical Hospital.

Through phone calls, participants responded to a questionnaire about adherence to daily vitamin/mineral supplementation ^7^, binge eating ^8^, depression symptoms ^9^, anxiety ^10^, health-related quality of life ^11^, physical activity level ^12^, and socioeconomic status ^13^.

Data are descriptively presented as mean (95%CI) or absolute and relative frequency (n, %). Individuals were categorically classified for the following parameters: adherence to daily vitamin/mineral supplementation (‘yes’ or ‘no’); binge eating score (≤ 17: ‘absence’, 18 – 26: ‘mild to moderate’, and > 27: ‘severe’); depression symptoms (score of 0 – 9: ‘no or minimal symptoms’, 10 – 18: ‘mild-to-moderate symptoms’, 19 – 29: ‘moderate-to-severe, 30 – 63: ‘severe symptoms’); and anxiety (score of 0 – 7: ‘normal or absence’, 8 – 15: ‘mild to moderate symptoms’, 16 – 25: ‘moderate to severe symptoms’, 26 – 63: ‘severe’). In addition, physically active and inactive patients were compared for health-related quality of life, depression and anxiety scores by Mann-Whitney test, using SPSS (Version 20.0, Chicago, IL). Significance level was set at p < 0.05.

Patients were 47.4 years old (44.6, 50.1), 56 (84.8%) were women, and 4 (8.7%) were smokers. Fifty-seven (86.3%) patients were economically vulnerable, with a self-reported family income < 485 BRL per month. Forty-eight (72.7%) and 18 (27.3%) underwent gastrointestinal bypass Y-in-Roux surgery and sleeve gastrectomy, respectively. The post-operative average time was 7.2 months (6.7, 7.9) and BMI was 35.5 kg/m^2^ (33.6, 37.4). Twenty-eight (60.9%) patients self-reported the presence of comorbidities. Patients self-reported the use of antihypertensive (15, 34.1%), antidiabetic (5, 11.4%), antihypothyroid (5, 11.4%), and antidepressant drugs (1, 2.3%).

Fifteen (30.0%), 11 (22.0%) and 15 (30.0%) patients were classified as having obesity grade I, II and III, respectively. Forty-nine (94.2%) had above reference waist circumference values. Thirty-four (65.4%) displayed high blood pressure. Twenty-three (16.7%) showed high glucose levels. Four (8.0%) had abnormal levels of HDL, LDL, and TG, whereas 12 (24%) showed CRP levels above the normal range (Figure 1, Panel A).

**Figure 1.**
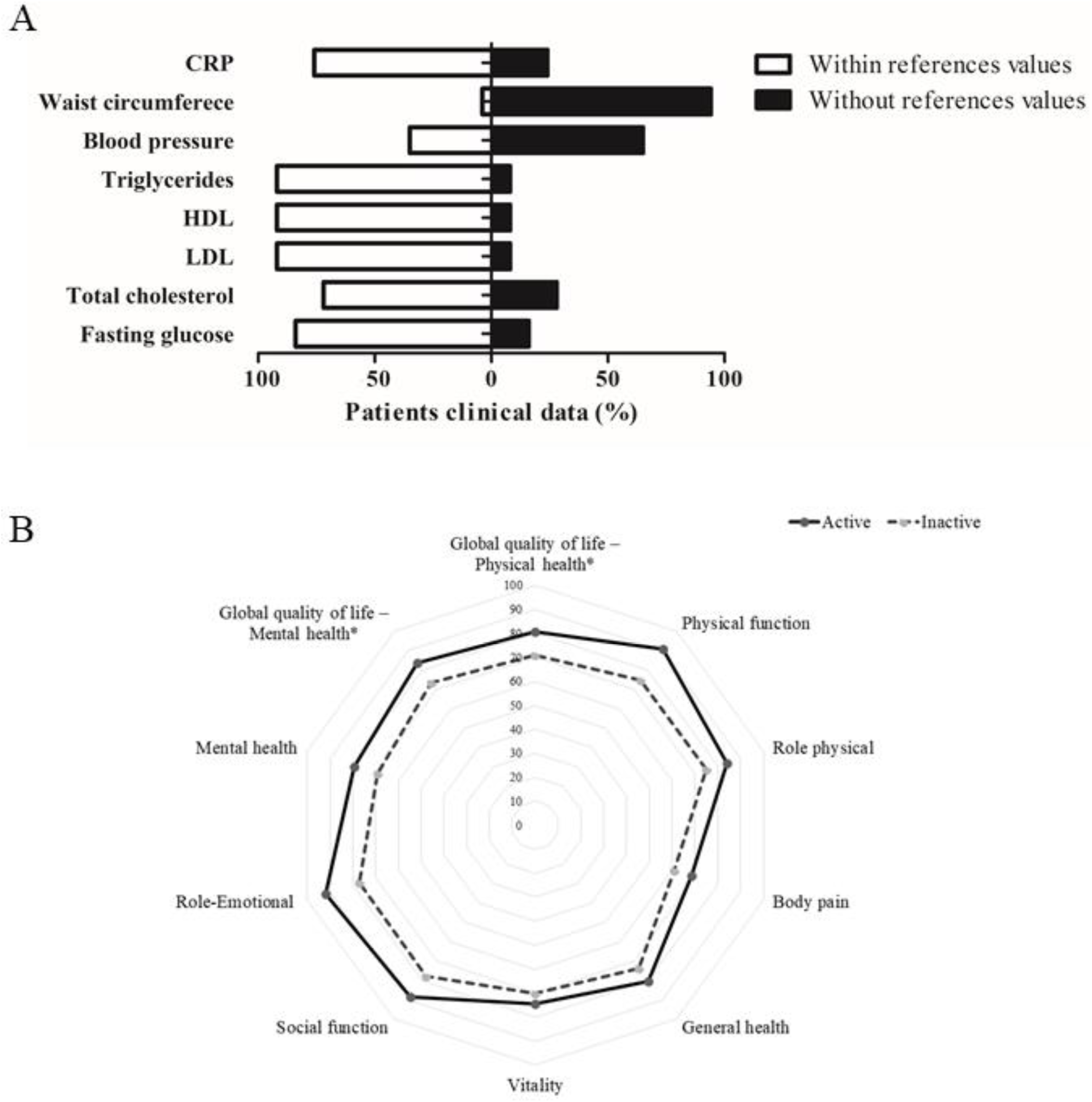
Panel A – Percentages of patients presenting with cardiometabolic risk factors. Panel B – Comparison between active and inactive patients for health-related quality of life. *p<0.05 between active and inactive.

Twelve (19.0%) patients reported irregular use of nutritional supplementation, and 4 (6.0%) patients reported binge eating habits.

Twelve (19.7%), 6 (9.8%), 2 (3.3%) exhibited mild-to-moderate, moderate-to-severe or severe symptoms of depression, respectively. Mild-to-moderate anxiety and moderate-to-severe or severe anxiety symptoms were reported by 17 (27.4%) and 7 (11.3%) of the patients, respectively. Three (4.5%) patients were urgently referred to a medical specialist because they exhibited suicidal ideation.

SF-36 physical score was 74.7 (70.2, 79.3) and mental health score was 77.4 (73.5, 81.4). Forty (59.6%) patients were classified as inactive. Of relevance, inactive patients showed worsen global scores of mental and physical health as compared with active peers (p = 0.02 and p = 0.04, respectively; Figure 1, Panel B). There were no differences between active and inactive for depression symptoms and anxiety scores (both p > 0.05).

The shortage of essential medical care has become a major health concern as the Covid-19 pandemic continues to unfold. The population of Sao Paulo, Brazil, has been severely impacted due to the very-high prevalence of Covid-19 cases, and as the health system promptly adjusted to accommodate infected patients, the problem of access to essential care escalated, leading to concerns about suboptimal care of patients with other chronic conditions. Patients who have had bariatric surgery are particularly vulnerable because they require monitoring for adverse surgical events and some are at increased risk for mental and physical disorders ^14, 15^. The Diabetes Surgery Summit consensus recently pointed out the importance of postoperative monitoring for this population in times of pandemic ^16^. Therefore, we screened recently operated patients in order to obtain objective data for overall health, quality of life and lifestyle. These patients were economically vulnerable, refrained from in-hospital care and advised to adhere to social isolation.

Our main findings suggest that a concerningly large percentage of patients presented with abnormal blood pressure, and ∼25% had increased systemic inflammation (assessed by CRP). Moreover, about one third showed mild to severe symptoms of depression, whereas ∼40% showed mild to severe anxiety symptoms. This suggests possible deviations from the lifestyle recommendations for postoperative care.

Approximately 20% of the patients were not complying with vitamin/mineral supplementation. Importantly, ∼60% failed to meet the prescribed physical activities recommendations, which is actually the reversed depiction of what we have observed in a group of patients from the same outpatient clinic before the pandemic (unpublished data). Sub-optimal nutritional care combined with physical inactivity may reverse the clinical improvements brought about by surgery, even in a short period of time ^17-19^. Of relevance, physically inactive patients exhibited lower mental and physical quality of life scores when compared with active ones, supporting the idea that lifestyle plays a role in the patients’ perceived well-being.

Overall, our data suggest that surveillance of patients who have undergone bariatric surgery during the current pandemic identifies cases needing direct face-to-face care with health providers. As a point of emphasis, three patients who revealed suicidal ideation had to be immediately referred to a specialist. Telemedicine strategies that are supervised by specialist bariatric and metabolic surgery providers have been recommended for management of patients who have had surgery. In deprived regions with limited or no access to telemedicine, ‘*in loco’* strategies may be the only viable alternative. In Brazil, the universal healthcare system works with ‘community health agents’, who are non-medical personnel that could be trained to help in the surveillance of at-risk groups, such as postoperative bariatric patients, during the pandemic.

The limitations of this study include the limited number of patients, the cross-sectional nature of the screening, and the need for more in-depth clinical assessments.

In conclusion, this out-of-hospital screening revealed that the absence of in-person health care due to the Covid-19 pandemic is associated with the emergence of moderate to severe physical and mental health conditions as well as lifestyle inadequacies among patients who have recently undergone bariatric surgery. The design of this study does not allow us to conclude whether the patients’ overall health deteriorated as a consequence of the pandemic, but it does indicate that there are a considerable number of post-surgery patients in need of direct health care.

## Data Availability

The authors will make the data available upon reasonable request.

## ACKNOWLEDGEMENTS

Sao Paulo Research Foundation – FAPESP (grants #2015/26937-4, #2017/13552-2, #2019/18039-7) and National Council for Scientific and Technological Development – CNPq (grant #402123/2020-4).

---

**Table 1.** Adherence to vitamin/mineral supplementation, physical activity levels, mood disorder symptoms, and quality of life.

| **Outcome** | **n (%) or mean (95%CI)** |
| --- | --- |
| **Adherence to daily vitamin/mineral supplementation (n=63)** |  |
| Yes | 51 (81.0%) |
| No | 12 (19.0%) |
| **Binge eating (n=66)** |  |
| Absence | 62 (93.9%) |
| Moderate | 3 (4.5%) |
| Severe | 1 (1.5%) |
| **Physical activity (n=66)** |  |
| Active | 26 (39.4%) |
| Inactive | 40 (60.6%) |
| **Depression symptoms (n=61)** |  |
| No or minimal symptoms | 41 (67.2%) |
| Mild-to-moderate | 12 (19.7%) |
| Moderate-to-severe | 6 (9.8%) |
| Severe | 2 (3.3%) |
| **Anxiety (n=62)** |  |
| Normal or absence | 38 (61.3%) |
| Mild-to-moderate anxiety | 17 (27.4%) |
| Moderate-to-severe | 7 (11.3%) |
| **Quality of life (n= 65)** |  |
| Global quality of life – physical health | 74.7 (70.2, 79.3) |
| Global quality of life – mental health | 77.4 (73.5, 81.4) |
| Physical function | 81.0 (75.7, 86.0) |
| Role physical | 78.0 (70.3, 86.5) |
| Body pain | 63.7 (57.0, 71.0) |
| General health | 76.1 (71.7, 80.5) |
| Vitality | 71.7 (66.8, 76.7) |
| Social function | 81.7 (76.2, 87.3) |
| Role emotional | 82.8 (75.8, 89.7) |
| Mental health | 73.1 (68.7, 77.5) |

